# Prion protein gene mutation detection using long-read Nanopore sequencing

**DOI:** 10.1101/2022.03.06.22271294

**Authors:** François Kroll, Athanasios Dimitriadis, Tracy Campbell, Lee Darwent, John Collinge, Simon Mead, Emmanuelle Vire

## Abstract

Prion diseases are fatal neurodegenerative conditions that affect humans and animals. Rapid and accurate sequencing of the prion gene *PRNP* is paramount to human prion disease diagnosis and for animal surveillance programmes. Current methods for *PRNP* genotyping involve sequencing of small fragments within the protein-coding region. The contribution of variants in the non-coding regions of *PRNP* including large structural changes is poorly understood. Here we use long-range PCR and Nanopore sequencing to sequence the full length of *PRNP*, including its regulatory region, in 25 samples from blood and brain of individuals with various prion diseases. Nanopore sequencing detected the same variants as identified by Sanger sequencing, including repeat expansions/contractions. Nanopore identifies additional single-nucleotide variants in the non-coding regions of *PRNP*, but no novel structural variants were discovered. Finally, we explore somatic mosaicism of *PRNP*’s octapeptide repeat region, which is a hypothetical cause of sporadic prion disease. While we find changes consistent with somatic mutations, we demonstrate that they may have been generated by the PCR. Our study illustrates the accuracy of Nanopore sequencing for rapid and field prion disease diagnosis and highlights the need for single-molecule sequencing methods for the detection of somatic mutations.

## Introduction

Prion diseases are a group of fatal mammalian neurodegenerative disorders, the most common of which in human is sporadic Creutzfeldt-Jakob disease (CJD). Prion diseases can be acquired or transmitted between humans and animals, necessitating active surveillance programmes. The infectious agent of prion disease is composed of assemblies of misfolded forms of cellular prion protein (PrP). Mutations in the gene that encodes PrP (*PRNP*) are associated with inherited prion diseases, which comprise a heterogeneous range of clinical phenotypes that mimic common dementias(1).

Human *PRNP* is composed of two exons flanking a single large intron for a total of 15 kb located on chromosome 20(2, 3). The 762-bp open reading frame is contained entirely within the second exon. Sanger sequencing of the protein-coding region of *PRNP* is routinely performed as part of the clinical investigation of patients suspected to have any form of prion disease(1). The coding region has been extensively studied in prion disease cases with over 40 pathogenic variants reported, most of which are non-synonymous single-nucleotide variants (SNV) (1). An important SNV in *PRNP* is at codon 129 (rs1799990), which encodes either methionine (M) or valine (V). Homozygosity at codon 129 is associated with an elevated risk of developing sporadic CJD and shorter survival relative to heterozygotes(4), and the variant is a strong determinant of clinical phenotype(5).

The N-terminal region of PrP contains a repetitive sequence of amino acids termed octapeptide repeat region (OPR). The normal OPR consists of a nonapeptide followed by four octapeptide repeats(6). Repeat length is variable in the human population ranging from deletions (OPRD, for octapeptide repeat deletion) of two repeats to insertions (OPRI, for octapeptide repeat insertion) of an extra 12 repeats. Insertion of four or more supplementary repeats is recognised as definite causes in inherited cases(1). Instability of the OPR, which might manifest as somatic insertion and/or deletion of repeats in the OPR in brain tissue, is a hypothetical mechanism of sporadic CJD(7-9).

The expression level of *Prnp* is a strong determinant of incubation time in mouse models of prion disease(10, 11). Variants in non-coding regions could lead to alterations in prion expression and thereby modify risk and impact on clinical features. In humans, SNVs in the intron or regulatory regions of *PRNP* might increase risk of sporadic CJD, potentially by raising *PRNP* expression (12-14). In cattle, a 23-bp deletion in the promoter and a 12-bp deletion in the intron increases an animal’s risk of developing bovine spongiform encephalopathy upon consumption of infected feedstuffs(15-17). As yet there has been no assessment of the non-coding regions of *PRNP* in CJD cases for structural variants that might alter regulatory regions.

Here we set up a long-read sequencing protocol for full-length human *PRNP*, including its regulatory region, in CJD patients. We used Oxford Nanopore MinION, a portable sequencer which offers a more complete view of genomic variation than short-read technologies as it generates long reads spanning repetitive regions and potential SVs(18, 19). Our aim was to establish a protocol that could potentially support (1) the rapid field sequencing of *PRNP*, (2) longer read lengths that are more likely to span a SV and can be more confidently aligned or assembled, and (3) high sequencing coverage to detect hypothetical somatic mutations in the

## Results

### Genotyping *PRNP* in prion disease cases using Nanopore sequencing

*PRNP* genotyping in prion diseases cases is routinely performed using both Sanger sequencing and gel electrophoresis of the protein-coding region(20). First, we tested if Nanopore sequencing performs as well as Sanger for the amplicon that is routinely used for clinical genotyping. Genomic DNA was extracted from CJD patient blood and the *PRNP* protein-coding region was amplified and sequenced using both Nanopore and Sanger. We computed a consensus sequence of the Nanopore reads and aligned it to the sequence obtained by Sanger sequencing. Both sequences were 100% identical. The sample was genotyped as codon 129 heterozygous (M129V) by both Sanger and Nanopore sequencing (Supplementary Fig. S1). This pilot experiment encouraged us to pursue Nanopore sequencing as an alternative to Sanger sequencing for *PRNP* genotyping.

Next, we developed a protocol for amplification and sequencing of the entire *PRNP* genomic region. We mapped the regulatory region based on genome annotations of epigenetic marks H3K4me1, H3K4me3, H3K27ac, and transcription factor binding sites(21). These pointed towards an important regulatory region starting around 1.1 kb upstream of exon 1 and extending into the intron. Although Nanopore allows sequencing of fragments longer than 1 Mb(22), we faced technical difficulties amplifying the 16.5 kb region as a single PCR product. This is likely due to a GC-rich (> 70%) region around exon 1. We therefore opted to sequence two overlapping amplicons (Fig. 1a). The smaller amplicon (2,988 bp) started upstream of the regulatory region and ended 1 kb into the intron. The larger amplicon (14,025 bp) included the remaining of the intron and exon 2. The OPR is composed of 5 similar repeats for a total of 123 bp (Fig. 1b).

**Figure 1.**
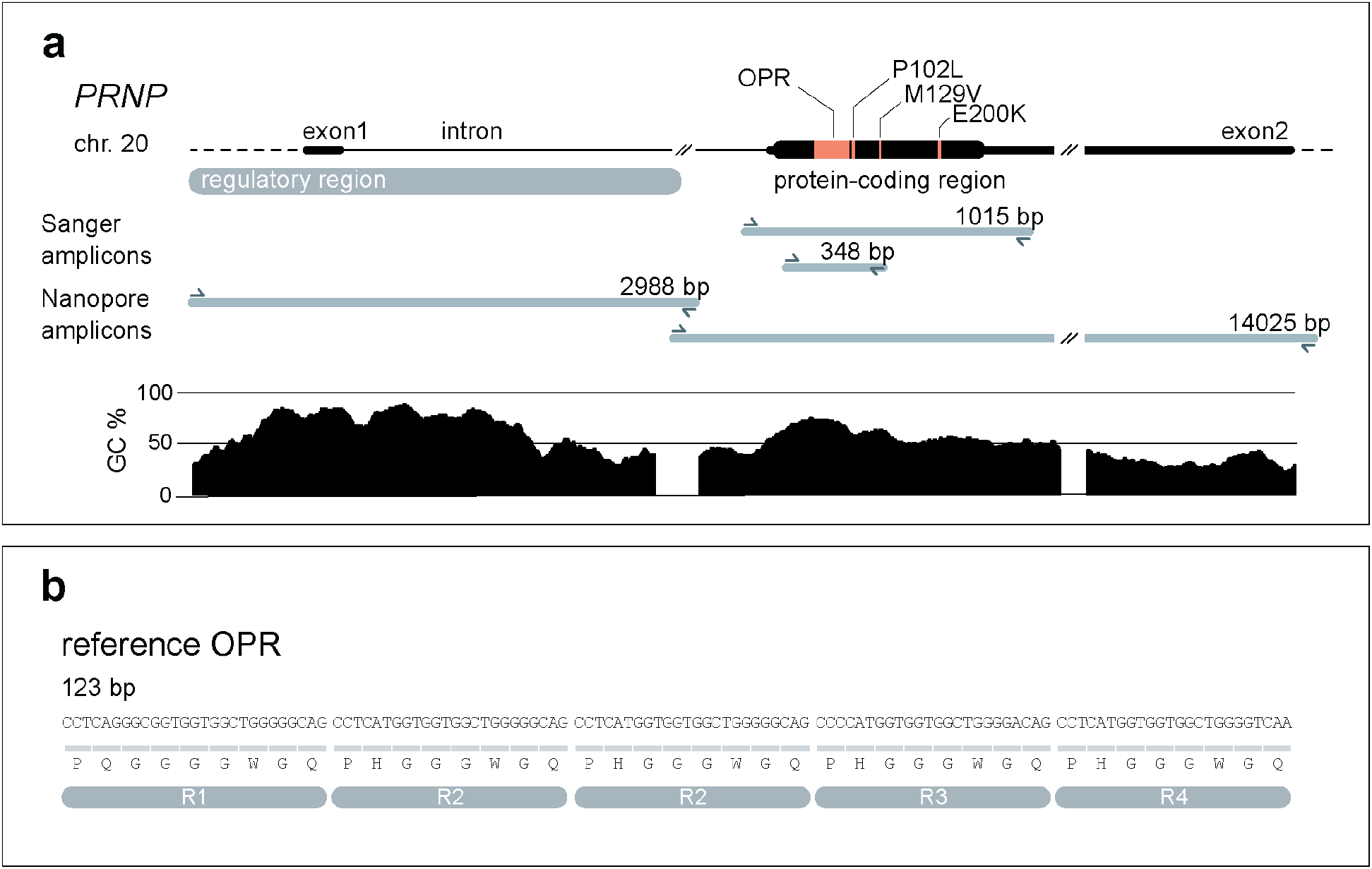
Nanopore sequencing of *PRNP* and octapeptide repeat genotyping. (**a**) The human *PRNP* gene. Important pathogenic variants, such as deletions/insertions in the octapeptide repeat region (OPR), codon 129 (M129V) or codon 200 (E200K) SNVs are indicated in the protein-coding region in orange. Genetic diagnosis at the MRC Prion Unit at UCL is performed by Sanger sequencing of the protein-coding sequence (1015-bp amplicon) or specifically of the OPR (348-bp amplicon). Exon 1 and the beginning of the intron are particularly GC-rich (see GC % plot). This likely explains why producing a single amplicon spanning the entire genomic region was technically challenging. Instead, we amplified *PRNP* in two overlapping fragments for Nanopore sequencing (see Nanopore amplicons). (**b**) Sequence of *PRNP*’s OPR. The OPR is composed of 5 similar repeats for a total of 123 bp. The first repeat (R1) is 27 bp and encodes a sequence of 9 amino acids (nonapeptide), the subsequent ones (R2, R2, R3, R4) are 24 bp and all encode the same sequence of 8 amino acids (octapeptide). The two R2 repeats are identical. R2 vary with R3 and R4 by two nucleotides at wobble positions.

Using this protocol, we sequenced the *PRNP* genomic region in 25 samples from 21 individuals. First, we sequenced one healthy control and seven patients with inherited prion disease—of which six carried an insertion and/or a deletion in the OPR (2 OPRD, 1 OPRD, 1 OPRI, 2 OPRI, 5 OPRI/1 OPRD, and 6 OPRI). Second, we sequenced both blood and brain samples from four inherited prion disease patients with OPR insertions. Third, we sequenced one healthy control and eight patients with sporadic CJD. Of the 21 individuals, nine carried a SNV in the protein-coding region detected by Sanger sequencing: one E200K, one P102L, and seven M129V (Table 1). Using Nanopore, we obtained a range of coverage of 3165– 8160× for the gene body (14,025-bp amplicon) and 133–8966× for the regulatory region (2,988-bp amplicon).

**Table 1.**

| ind. | sample | tissue | prion disease | Sanger sequencing |  | Nanopore MinION |  |  |
| --- | --- | --- | --- | --- | --- | --- | --- | --- |
|  |  |  |  | SVs | SNVs | SVs | SNVs (protein-coding) | non-coding SNVs |
| 1 | 52331 | blood | control | - | not tested | - | M129V | 14 |
| 2 | 1906 | blood | inherited | 2 OPRD | - | 2 OPRD | - | 3 |
| 3 | 55826 | blood | inherited | 1 OPRD | - | 1 OPRD | - | 9 |
| 4 | 58398 | blood | inherited | - | E200K | - | E200K | 1 |
| 5 | 58778 | blood | inherited | 1 OPRI | - | 1 OPRI | P102L | 4 |
| 6 | 57749 | blood | inherited | 2 OPRI | M129V | 2 OPRI | M129V | 17 |
| 7 | 46345 | blood | inherited | 5 OPRI/1 OPRD | - | 5 OPRD/1 OPRD | - | 5 |
| 8 | 58648 | blood | inherited | 6 OPRI | - | 6 OPRI | - | 4 |
| 9 | 43706 | brain | inherited | 4 OPRI | - | 4 OPRI | - | 1 |
| 9 | 42656 | blood | inherited | 4 OPRI | - | 4 OPRI | - | 1 |
| 10 | 57265 | brain | inherited | 4 OPRI | M129V | 4 OPRI | M129V | 13 |
| 10 | 55492 | blood | inherited | 4 OPRI | M129V | 4 OPRI | M129V | 13 |
| 11 | 59060 | brain | inherited | 6 OPRI | - | 6 OPRI | - | - |
| 11 | 53747 | blood | inherited | 6 OPRI | - | 6 OPRI | - | - |
| 12 | 53689 | brain | inherited | 8 OPRI | M129V | 8 OPRI | M129V | 15 |
| 12 | 47875 | blood | inherited | 8 OPRI | M129V | 8 OPRI | M129V | 15 |
| 13 | 56635 | blood | control | - | M129V | - | M129V | 16 |
| 14 | 54493 | blood | sporadic | - | M129V | - | M129V | 17 |
| 15 | 54890 | blood | sporadic | - | - | - | - | - |
| 16 | 54917 | blood | sporadic | - | - | - | - | 4 |
| 17 | 54960 | blood | sporadic | - | M129V | - | - | 16 |
| 18 | 54968 | blood | sporadic | - | - | - | - | 1 |
| 19 | 55048 | blood | sporadic | - | - | - | - | 1 |
| 20 | 55050 | blood | sporadic | - | - | - | - | - |
| 21 | 55052 | blood | sporadic | - | M129V | - | M129V | 14 |

First, we called SNVs for all samples. After filtering calls based on allele frequency and strand bias, we detected a total of 44 different SNVs, most of which were present in more than one individual, for a total of 198 calls (Fig. 2a, Supplementary Fig. S2). All 44 SNVs were found in human genetics databases, and their respective frequency in the general population was in accordance with the number of individuals carrying each SNV in our panel (Fig. 2b). The SNV calling algorithm successfully called the E200K, P102L, and M129V variants previously identified by Sanger sequencing (Table 1). No additional SNVs were detected in the protein-coding sequence, indicating a low frequency of false positive calls. We did not find an excess of SNVs in sporadic CJD cases (Supplementary Fig. S2).

**Figure 2.**
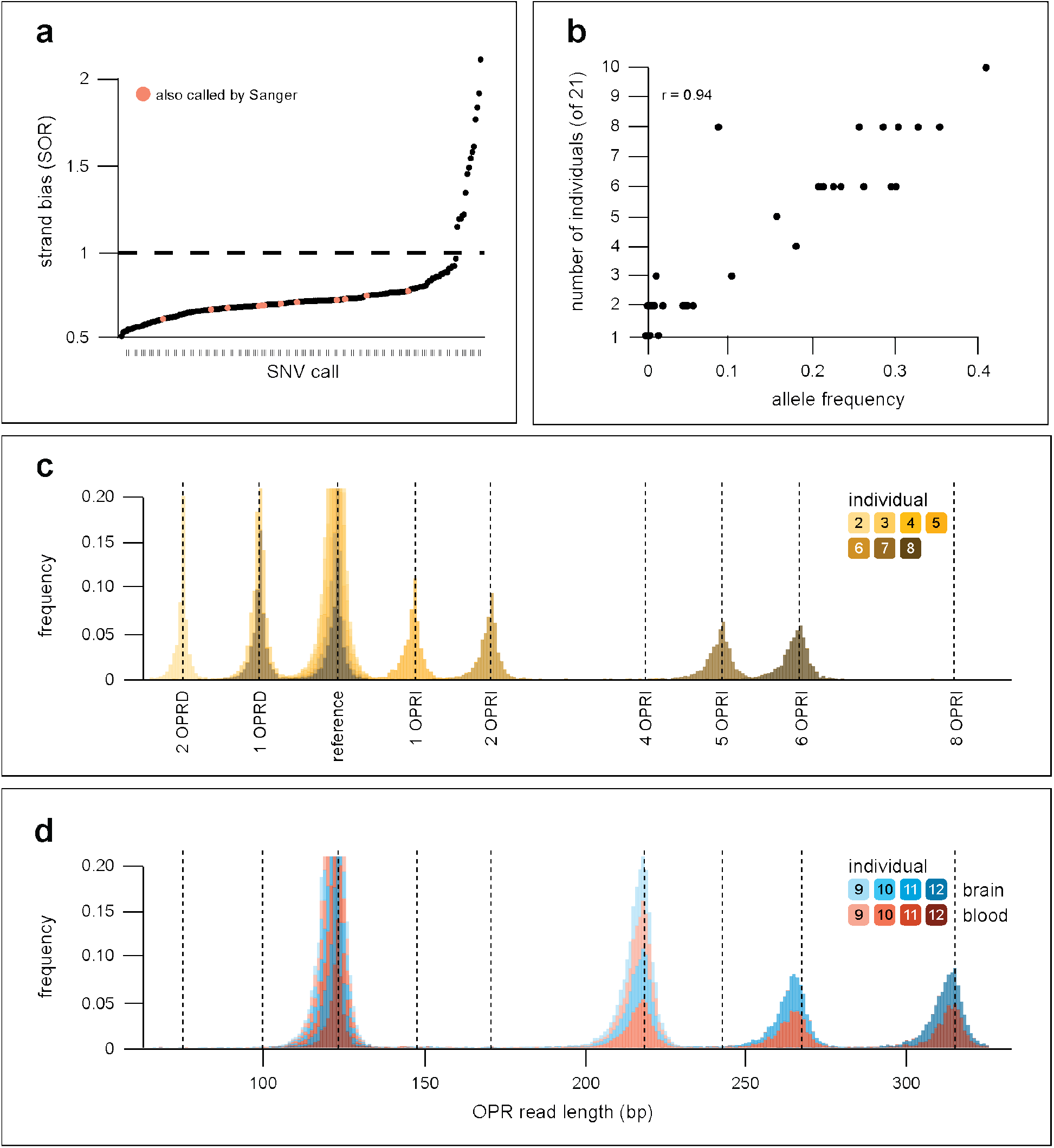
Detection of known variants in *PRNP* with Nanopore sequencing. (**a**) Each dot represents a SNV call by Nanopolish. SNV calls are ranked by increasing evidence of strand bias (SOR, strand odds ratio). In orange are the SNVs in the protein-coding region which were also called by Sanger. They are together the set of true positives we used to calibrate our strand bias threshold (SOR = 1.0, dashed black line). (**b**) Number of individuals from our panel who carried each unique SNV as a function of allele frequency in the general population. r = 0.94 by Pearson’s correlation. (**c**) Lengths of the OPR reads in seven inherited CJD cases. (**d**) Lengths of the OPR reads in four inherited cases for which genomic DNA from blood and brain was sequenced. See Table 1 for more details about individuals and samples.

Second, we called SVs for all samples. The variant calling algorithm correctly identified all 15 OPR mutations present in our panel including the precise size of the inserted/deleted sequence ± 2 bp (Fig. 2c–d, Table 1). We did not detect novel SVs in *PRNP*’s regulatory region or in the sporadic CJD samples. These results support the use of Nanopore sequencing for *PRNP* genotyping in patient samples.

### Exploring somatic mutation of the octapeptide repeat region using Nanopore sequencing

Somatic mosaicism has been implicated in many diseases, from cancer to neurodegeneration (23-26). In prion diseases, somatic mutation of *PRNP*’s OPR has been hypothesised as a possible cause of sporadic CJD(8, 9). We took advantage of the high sequencing coverage obtained with Nanopore sequencing to search for rare somatic insertions or deletions in *PRNP*’s OPR.

We first trimmed all reads we collected to keep only *PRNP*’s OPR, generating 208,554 OPR reads. Then, we labelled each read with the most likely OPR length based on the total insertion and/or deletion compared to the reference. We designed a consensus sequence for one OPR repeat and built a set of template sequences for a range of possible OPRs. Each read was then aligned to the OPR template sequence matching its OPR label, returning the number of mismatches. We defined a somatic mutation call as any read whose OPR label was not reference or the genotype of its sample and was at least 94% identical to its OPR template sequence (Supplementary Fig. S3). For example, any OPR read with a 24-bp insertion was labelled 1 OPRI, aligned to the 1 OPRI template, and was selected as a somatic mutation call if it aligned to its template sequence and did not come from a sample with a heterozygous 1 OPRI mutation.

Chimeric reads, which are single reads composed of sequences from one or more amplicons, can occur with Nanopore sequencing(27). As we sequenced samples with different OPR genotypes on the same flow cell, we wanted to exclude any somatic mutation call which could be a chimeric read from another sample on the flow cell. To this end, we discarded the following reads: those not barcoded at both ends; those containing an adapter or barcode in their middle; those longer than their amplicon(28). We identified a total of 129 somatic mutation calls, at a frequency per sample of 0–0.28% of reads (Fig. 3a, Supplementary Fig. S5). Reads from the forward and reverse strand were uniformly represented in the somatic mutation calls, excluding the possibility of strand bias. The number of somatic mutation calls from each sample did not correlate with its sequencing coverage, which likely excludes sequencing errors occurring at the pore (Supplementary Fig. S4)(29). Based on the above checks, we concluded that the 129 somatic mutation calls are not sequencing artefacts.

**Figure 3.**
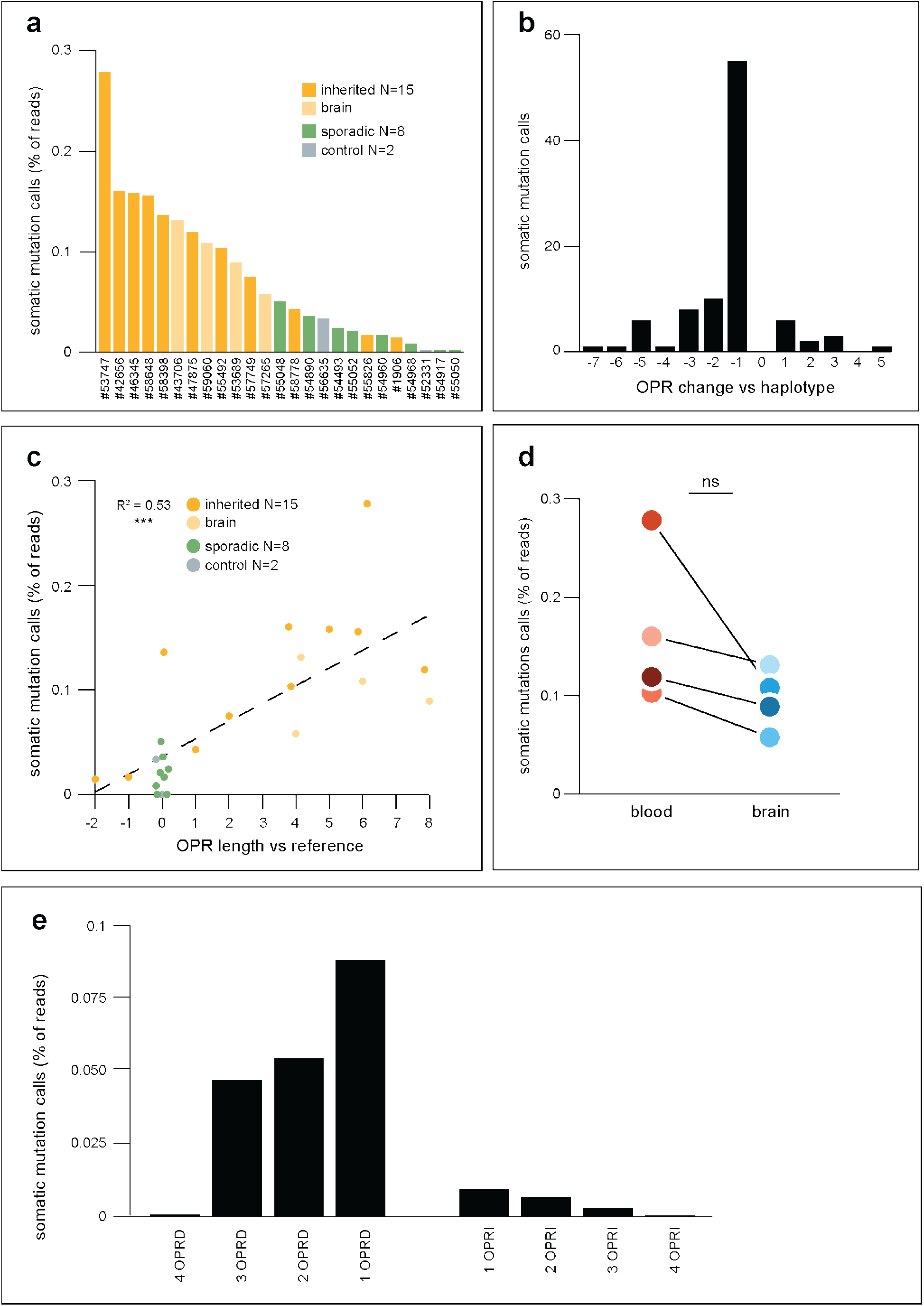
Somatic mutation calls in *PRNP*’s OPR. (**a**) Somatic mutation calls per sample, as percentage of the number of reads. (**b**) Number of somatic mutation calls as a function of the change it generated in the OPR. For example, a 3 OPRI somatic mutation call on the mutated OPR haplotype of a 4 OPRI sample was counted as a -1 change. Of the N = 94 somatic mutation calls from haplotype-phased samples, 55 (59%) removed one repeat from the OPR (−1 changes). (**c**) Somatic mutation calls, as percentage of reads of each sample, as a function of the OPR length of each sample. The length is given compared to the reference OPR, 0 corresponds to the reference OPR, negatives correspond to OPR deletions (e.g. -2 is 2 OPRD), and positives correspond to OPR insertions (e.g. 8 is 8 OPRI). The black dashed line is the line of best fit by linear regression: *% somatic mutation calls = 0*.*036 + 0*.*017 × OPR length*. OPR length is a significant predictor of the percentage of somatic mutation calls, R^2^ = 0.53, *** p < 0.001 by linear regression. (**d**) Somatic mutation calls, as percentage of reads of each sample, for four inherited CJD cases which had genomic DNA from both blood and brain sequenced. For each individual, there was a decrease in somatic mutation calls in brain compared to blood. p = 0.068 by one-sample one-sided t-test (null hypothesis was no decrease between blood and brain). (**e**) Changes in the OPR introduced by PCR amplification of a reference OPR, as in (a).

Of the 129 somatic mutation calls, 103 (80%) were from inherited CJD samples (Supplementary Fig. S3). This represented a significant excess of calls from this group compared to sporadic CJD or control samples, even after accounting for the higher number of inherited CJD samples in our panel. Most inherited CJD patients in our panel carried a monoallelic OPR mutation. In these samples, the somatic mutation calls could represent the mutation of the reference allele or the mutation of the allele already carrying an OPR mutation. For example, a 5 OPRI somatic mutation call in a 4 OPRI heterozygous carrier could represent a 5-repeat insertion in the reference allele, or a 1-repeat insertion in the 4 OPRI allele. We used haplotype phasing to discriminate between these two possibilities (Supplementary Fig. S4). Of the 64 somatic mutation calls from samples with an OPR mutation, 47 (73%) were assigned to the OPR-mutated allele (Supplementary Fig. S3). Somatic mutation calls ranged from the deletion of 7 OPR repeats to the insertion of 5 additional repeats, but the majority (55/94, 59% of haplotype-assigned calls) represented the deletion of one OPR repeat (Fig. 3b).

Were specific samples more likely to produce somatic mutation calls? Samples from individuals with longer OPRs had more somatic mutation calls, with each additional repeat in the OPR increasing somatic mutation calls by 0.017% (Fig. 3c). In the four individuals who had both blood and brain DNA sequenced, we observed a trend towards a lower frequency of somatic mutation calls in brain compared to blood (0.07 ± 0.07% fewer calls in brain vs blood of the same individual, Fig. 3d).

### PCR amplification introduces errors in the octapeptide repeat region that resemble somatic mutations

Li et al. have elegantly demonstrated that *PRNP*’s OPR is prone to either contraction or expansion when subjected to PCR amplification and when replicated in wild-type *E. coli* cells(8). Therefore, we tested if the somatic mutation calls could be errors generated by the *Taq* polymerase. We amplified the protein-coding region (1015-bp amplicon, Fig. 1a) from a known amount of control sample: 1 ng of genomic DNA (#56635), approximately 307 haploid genomes. In this sample, we previously found that 0.0325% of reads were somatic mutation calls. Assuming 0.0325% of genomes in the original genomic DNA carried a somatic mutation of the OPR, it was almost certain that we drew either no (0.9 probability) or only one mutated genome (0.09 probability, cumulatively 0.99 probability). Accordingly, if amplification did not introduce errors in the OPR, only reference OPR reads or a few mutated OPR reads with a unique mutation (e.g. all 2 OPRI) should be detectable in the sequencing reads. However, 0.21% of reads were somatic mutation calls, ranging from 4 OPRD to 4 OPRI (Fig. 3e). The distribution of OPR lengths resembled the distribution of the original 129 somatic mutation calls (Fig. 3e vs Fig. 3b) with 1 OPRD being the most frequent call (319/761 calls, 42%). Therefore, we cannot be certain the 129 calls are true somatic mutations Instead, the calls may have been errors introduced by the PCR. Single-molecule sequencing technologies are likely required to confirm the presence of somatic mutations in *PRNP*’s OPR in human samples.

## Discussion

Sequencing of *PRNP* is of crucial importance in informing patients, veterinaries, and clinicians about prion diseases diagnosis and management. Here we established a novel long-read sequencing strategy to sequence the full length of *PRNP* including its regulatory region (16.5 kb). First, we showed that Nanopore sequencing accurately detects known single-nucleotide and structural variants in *PRNP*, including the insertion and deletion of repeats in the OPR that are not straightforward to report with short read length Sanger sequencing. Second, we did not discover novel SVs in non-coding regions of *PRNP* or in patients with sporadic CJD. Third, we detected rare changes in the OPR consistent with somatic mutations, which have been speculated to be the cause of sporadic CJD. However, we demonstrated that these somatic mutation calls may in fact represent errors introduced during the PCR.

The present strategy has some limitations: it requires amplification of *PRNP* in two fragments, which renders the protocol relatively laborious and lab-based. Removing the amplification steps could help streamline the process. To do so, targeted sequencing using CRISPR-Cas9(30) or the ‘Read Until’ mode of Oxford Nanopore devices could be employed(31). By removing the amplification steps, these strategies could enable *PRNP* sequencing in the field. This could benefit rapid decision making during therapeutic studies and support programmes which survey the zoonotic potential of animal prion diseases, such as the recent episodes of Chronic Wasting Disease in Norway and North America(32, 33). Furthermore, direct sequencing of genomic DNA would allow the analysis of epigenetic marks, which may serve as a predictor of disease progression in sporadic CJD(30).

Somatic mutations in *PRNP* have been suspected to be a cause of sporadic CJD. The hypothesis posits that the first prion in a patient with sporadic CJD originated in a cell or group of cells which acquired somatic mutation in *PRNP*. A strong candidate for this original mutation is the somatic insertion of additional repeats in the OPR, as OPRIs can cause inherited CJD and somatic instability is typical for repetitive sequences(34). A possible mechanism for the insertion or deletion of repeats in the OPR is replication slippage(8, 35). During replication, the DNA polymerase may dissociate from the DNA leading to either the template or daughter strand to ‘slip’, i.e. re-anneal incorrectly to an earlier repeat (Supplementary Fig. S6). In the case of template strand slippage, the polymerase then skips one or more repeats when DNA replication resumes, producing an OPRD. In the case of daughter strand slippage, the DNA polymerase replicates the same one or more repeats again, which produces an OPRI. In our panel, most somatic mutation calls in inherited CJD samples were assigned to the OPR-mutated allele and OPR length positively correlated with frequency of somatic mutation calls. Both observations support this model: as more repeats are present in the DNA molecule undergoing replication, there are more ways the two DNA strands can mispair if the polymerase dissociates, leading to OPR mutations.

However, our results suggest that the OPR mutations identified here might have been generated by the *Taq* polymerase. The strong bias towards 1 OPRD in our calls also supports this possibility. Indeed, deletion of repeats are more frequent in bacteria, while eukaryotic repetitive sequences show no bias or a bias towards insertions(36). Replication slippage is dependent on cell division, which leads to two predictions relevant for future research on sporadic CJD. First, the frequency of somatic mutations of the OPR should be higher in cells undergoing divisions than in post-mitotic cells. In four individuals, we found that the frequency of somatic mutation calls was consistently higher in blood DNA than in brain DNA, which suggests that some calls could be genuine somatic mutations. Second, the original OPR mutation which causes sporadic CJD may not arise in post-mitotic neurons, but rather during development or in glia. Of note, mechanisms independent of cell division may also be possible, as has been suggested for trinucleotide repeat disorders such as Huntington disease(37, 38). Future research should initially aim at discovering genuine somatic mutations of *PRNP*’s OPR in genomic DNA from donors, for example using single-molecule sequencing technologies(39).

## Methods

### Patient recruitment and sample obtention

Healthy control donors were recruited from spouses or relatives of patients by the National Prion Clinic (London, UK), and the UCL Dementia Research Centre (London, UK). All experimental protocols were approved by the London Queen Square Research Ethics Committee (reference 05/Q0505/113). Samples were obtained with written informed consent from all controls, patients, or a patient’s consultee in accordance with applicable UK legislation and Codes of Practice. All methods were carried out in accordance with relevant guidelines and regulations.

### DNA extraction

Nucleon BACC3 kit was used for DNA extraction from frozen blood samples according to the manufacturer’s protocol. Final DNA samples were stored at 4°C until needed. DNA extraction from CJD brain samples was carried out in a Biosafety Level 3 facility as previously published(40).

### Sanger sequencing

Sanger sequencing of the protein-coding sequence of *PRNP* was performed following published protocols for genetic diagnosis(20, 41). The amplicon subjected to sequencing was the 1015-bp amplicon covering the protein-coding region (Fig. 1a). The shorter amplicon (348 bp in the reference allele) was used to assess OPR deletions/insertions in affected individuals by gel electrophoresis.

The Sanger sequencing trace included in Supplementary Fig. S1 was from Benchling (benchling.com).

### Nanopore sequencing of *PRNP*’s protein-coding region

PCR was performed on genomic DNA from blood of a healthy individual to produce the 1015-bp amplicon covering the protein-coding sequence (see above). Forward primer was 5’-CTATGCACTCATTCATTATG-3’, reverse primer was 5’-GTTTTCCAGTGCCCATCAGTG-3’. The PCR well contained 12.5 μL 2× MegaMix Royal (Microzone), 10.5 μL H2O, 1 μL primers (12.5 μM each), 1 μL genomic DNA. Initial denaturation was 5 min at 95°C, followed by 35 cycles of: 95°C for 30 sec (denaturation); 58°C for 40 sec (annealing); 72°C for 1 min (extension). The PCR product was purified and concentrated using Zymo DNA Clean & Concentrator-5 kit and eluted in 50 µL TE buffer. The PCR product’s concentration was quantified with Qubit (dsDNA Broad Range assay) and its length was verified using TapeStation 2200 (D1000 tape). Library preparation was performed according to Nanopore Technologies’ 1D amplicon by ligation protocol (version *ADE_9003_v108_revU_18Oct2016*). All reagents were provided by the SQK-LSK108 kit, except: NEBNext Ultra II End Repair/dA-Tailing buffer and enzyme mix (#E7546), NEB Blunt TA/Ligase Master Mix (#M0367), and Agencourt AMPure XP beads (#A63880). The PRNP coding region amplicons were diluted after end-repair/dA-tailing to bring only 0.2 pmol of DNA fragments to the ligation reaction. 23 ng of final library were loaded onto the MinION flow cell. Sequencing was performed for 53 minutes and followed live on the MinKNOW software.

Fast5 files were basecalled using guppy v4.5.3 (Oxford Nanopore Technologies). We then computed a consensus sequence of the Nanopore reads using canu v2.1.1(42), and “polished” it using nanopolish v0.13.2(43). The resulting consensus sequence was aligned to the sequence obtained by Sanger sequencing using BLASTn (megablast) (NCBI). The full alignment is included in Supplementary Fig. S1b.

To call variants, the reads obtained after basecalling were aligned to the human reference genome (hg38) using minimap2 v2.18-r1015(44). The resulting sam file was converted to bam, then sorted and indexed using samtools v1.9(45). Nanopolish v.0.13.2 was used for variant calling. Alignments were visualised with the Integrative Genomics Viewer (IGV) v2.8.6.

### Nanopore sequencing of full-length *PRNP* and its regulatory region

Amplification prior to Nanopore sequencing was performed in two subsequent PCRs. The first one amplified the genomic region of interest from genomic DNA, the second attached unique barcodes to allow multiplexing of several samples onto the sequencing flow cell. In order to use the barcoding primers provided by Oxford Nanopore Technologies in the PBK-004 barcoding kit, we designed primers that included a complementary sequence to the barcoding primers (see *Primer sequences*). We used the NEB LongAmp *Taq* DNA polymerase, which performed better than other long-range polymerases we tried.

The first PCR amplified *PRNP* in two amplicons (Fig. 1a, Nanopore amplicons). For the regulatory region amplicon (chr20:4,685,060–4,688,047), primers were primer 3 (forward) and primer 4 (reverse); annealing was 59°C – 25’’; elongation time was 2’40’’. For the gene body amplicon (chr20:4,685,060–4,701,756), primers were primer 2 (forward) and primer 1 (reverse); annealing was 57°C – 30’’; elongation time was 13’00’’. The reactions and other program settings were as recommended by the manufacturer’s protocol. Template DNA was 50–400 ng.

The barcoding PCR and library preparation were carried out according to ONT’s barcoding kit (SQK-PBK004, version PBK_9073_v1_revB_23May2018) and ligation sequencing kit (SQK-LSK109) protocols. Up to 12 samples were multiplexed on each flowcell and sequencing was performed for 48 hours.

### Basecalling and alignment

The raw, multi-line fast5 files were processed using ONT Guppy v4.2.2 suite. We used guppy_basecaller for basecalling and guppy_barcoder for demultiplexing (assigning reads to each samples). The reads in fastq format were then aligned to the human reference genome (hg38) using guppy_aligner. Samtools v1.7.0 was used to sort and index the resulting binary files (bam format). IGV v.2.8.6 was used to visualise the alignments.

### Single-nucleotide variant calling

We used nanopolish v0.13.3 to call single-nucleotide variants (SNVs) in the *PRNP* genomic window. Default nanopolish filtering criteria were used: only SNVs in regions sequenced at a minimum coverage of 20× and with an allele frequency of minimum 0.2 were called. We filtered the calls further using strand bias as a criterion(30). MinION sequences at random the forward or reverse strand. Hence, for a true positive variant call, the reads supporting the variant are predicted to be ∼ 50% forward and ∼ 50% reverse. There is evidence of strand bias if this proportion is strongly imbalanced, i.e. when the forward and the reverse reads do not uniformly call for the same nucleotide. We used the StrandOddsRatio (SOR) metric computed by nanopolish(46), which increases with evidence of strand bias. To filter SNV calls in non-coding regions of *PRNP*, we reasoned that the SNVs in the protein-coding region identified by both Sanger and Nanopore sequencing could be used as a set of true positives. For those, the maximum SOR was 0.83 (N = 12). Therefore, we filtered out any SNV call above SOR = 1.0 as showing evidence of strand bias and hence a possible false positive. This decreased the number of SNV calls for all samples (N = 25) from 275 to 198. SNV calls before and after strand-bias filtering are included (Additional File 1).

### Haplotype phasing

To assign the gene body reads to haplotypes (haplotagging), we generated a new VCF file for each file containing its filtered SNV calls (see *Single-nucleotide variant calling*). Five samples (#58648, #54890, #55050, #59060, #53747) had no heterozygous SNV call and therefore their reads could not be haplotagged. We first used whatshap phase to phase the SNV calls in relation to each other(47). A single SNV call is sufficient for haplotagging but could not be phased by whatshap phase. Therefore, the VCF files of samples with a single SNV call (#54917, #54968, #55048, #43706, #42656) were manually edited to match the formatting of a phased variant call. We then used whatshap haplotag to haplotag the reads.

### Structural variant calling

Prior to SV calling, alignments were filtered further to keep only high-quality reads. For the regulatory region amplicon, any read shorter than 1 kb, longer than 3.5 kb, or with more than 15% of its length soft-clipped was discarded. For the gene body amplicon, any read shorter than 10 kb, longer than 15 kb or with more than 5% of its length soft-clipped was discarded. The goal of the maximum length criteria is to exclude potentially chimeric reads. Secondary alignments were also discarded.

We used sniffles v1.0.12 to call structural variants (SVs) larger than 20 bp and supported by at least 100 reads(48). Calls were filtered to keep SV calls within the *PRNP* genomic window, with an allele frequency above 0.15 and not showing evidence of strand bias (StrandBias filter raised by sniffles).

### Lengths of OPR reads

This refers to Fig. 2c–d. The filtered aligned reads (see above) from the gene body were trimmed to keep only the OPR (chr20:4699371– 4699493) using samtools ampliconclip(45). From the reads we obtained, any read shorter than 21 bp or longer than 702 bp were discarded. 21 bp was chosen as minimum as it would correspond to a 4 OPRD, minus 6 bp to account for small artefactual deletions. 702 bp was chosen as maximum as it would correspond to a 24 OPRI, plus 3 bp to account for small artefactual insertions. We then counted the number of reads of each length to produce the histograms in Fig. 2c–d.

### Search for somatic mutations of the OPR

The haplotagged OPR reads were imported in R from SAM files, for a total of 208,554 OPR reads. We parsed the CIGAR of each read to calculate its total insertion and deletion. For example, 11500H43M1I24M1D29M1I7M3D7M2D7M2306H returned insertion 2 bp (1I + 1I) and deletion 6 bp (1D + 3D + 2D). The hard clips (11500H, 2306H) were created by samtools ampliconclip. Each read was then assigned to a most likely OPR genotype based on its total insertion/deletion. For each OPR genotype, the interval of possible lengths was defined as the target insertion/deletion length minus 6 bp to allow small artefactual deletions up to the target insertion/deletion length plus 3 bp to allow small artefactual insertions. For example, the target deletion length for 1 OPRD is -24 bp. Therefore, any read with a total deletion of -30 bp up to -21bp was labelled as a potential 1 OPRD read. As the target insertion/deletion for the reference OPR is 0 bp, the interval was set from deletions of -6 bp up to insertions of 3 bp. Any read with a total insertion/deletion not included in one of the intervals (e.g. a 10-bp insertion) was labelled as unassigned.

At this stage, we generated Supplementary Fig. S5 by plotting for each sample the proportion of reads labelled as reference or as the expected OPR mutation assigned to each haplotype.

In parallel, we built OPR template sequences using the OPR consensus sequence. Each read, except if labelled as unassigned, was then aligned one-to-one to the OPR template matching its OPR label (Supplementary Fig. S5), returning the number of mismatches. Alignments were performed with ClustalW(49), implemented in the msa R package(50). The mismatch threshold to filter somatic mutation calls was calculated as the mean + standard deviation of the number of mismatches of all ‘expected’ reads, i.e. reference reads and reads matching the OPR genotype of their sample. Somatic mutations calls were then defined as any read not labelled as reference (Supplementary Fig. S5), not labelled as the OPR genotype of its sample, and having fewer mismatches with its template than 5.8% of its length. For the compound heterozygous sample #46345, reads labelled as reference and below the mismatch threshold were also defined as somatic mutation calls.

### Control PCR to test for PCR-introduced errors

1 ng of genomic DNA from sample #56635, approximately 307 haploid genomes(51), was PCR amplified to produce the 1015-bp amplicon covering the protein-coding sequence (see *Sanger sequencing*). The product was cleaned, eluted in 25 µL of dH2O, and its concentration was quantified at 10.36 ng/µL by Qubit (dsDNA Broad Range assay). 1 µL of this product was then barcoded and sequenced as previously (see *Nanopore sequencing of full-length PRNP […]*).

The fast5 files were basecalled to produce reads which were aligned to the human reference genome as previously (see *Basecalling and alignment*). We then filtered the reads by discarding any read shorter than 900 bp, longer than 1300 bp, or with more than 20% of its length soft-clipped. We trimmed the reads to keep only the OPR (see *Lengths of OPR reads*), generating 366,243 OPR reads. Calling possible somatic mutations was then performed as previously (see *Search for somatic mutations of the OPR*).

The original frequency of somatic mutation calls in sample #56635 was 0.0325% (9 of 26,764 OPR reads were somatic mutation calls). Let us assume that these somatic mutation calls were genuine, and that any genome taken at random from sample #56635 had a probability of carrying an OPR mutation *P*(*mutated*) = 0.000325. Conversely, the probability that any genome taken at random from sample #56635 was OPR reference was *P*(*reference*) = 1 - 0.000325 = 0.99967.

The probability to have drawn exactly 0 genome of the 307 can be written as the probability that all 307 genomes were reference, which is *P*(*reference*)^307^ = 0.99967^307^ = 0.904, i.e. there was a 90.4% probability to draw 0 mutated genome in the 307.

The probability that exactly *n* genomes out of *k* genomes drawn (here, 307) were mutated can be calculated as

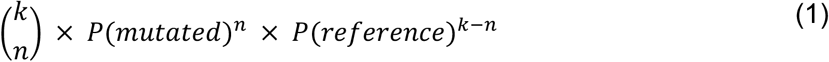

where

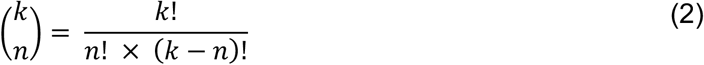

From Equation 1 and 2, the probability of having drawn exactly 1 mutated genome in the 307 was 307 × 0.000325^1^ × 0.99967^307−1^ = 0.0902, i.e. there was a 9.02% probability of drawing exactly 1 mutated genome in the 307.

The probability of drawing 0 or 1 mutated genome in the 307 was therefore 0.904 + 0.0902 = 0.9942, i.e. the probability to draw more than 1 mutated genome was 0.58%.

### Primer sequences

The sequence complementary to the ONT barcoding primers (kit PBK-004) is shown in bold.

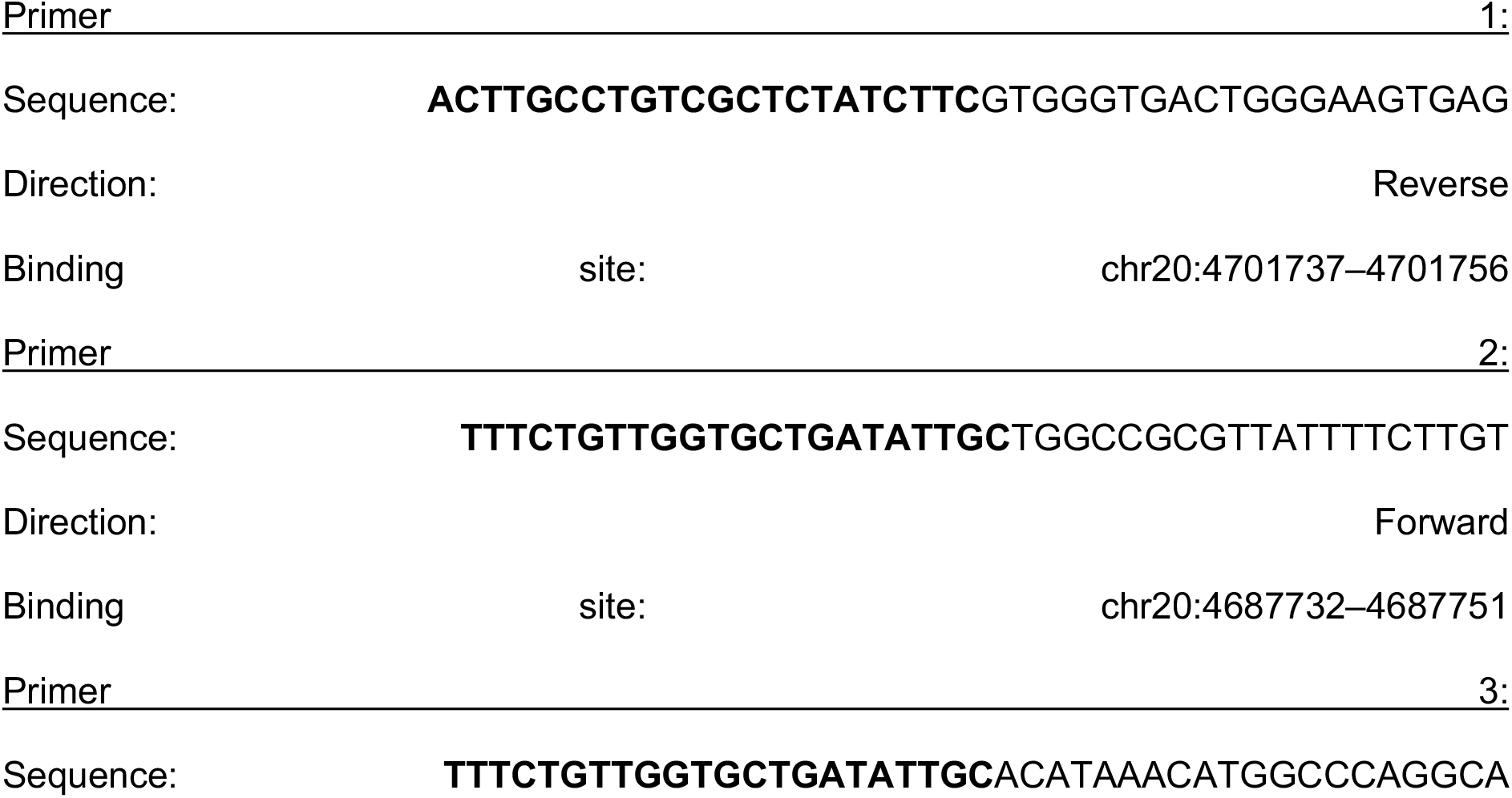
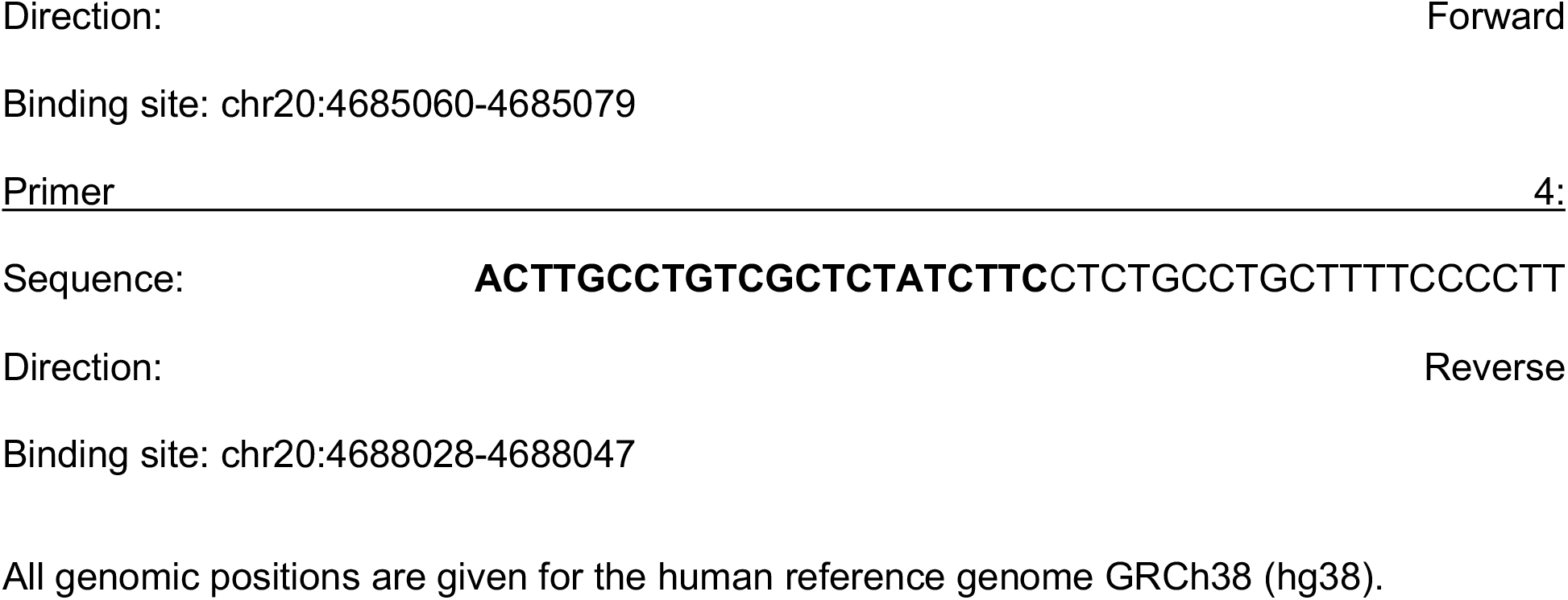

## Data Availability

Code will be made available at https://github.com/athanadd/prnp-nanopore-seq and https://github.com/francoiskroll/prnp_nanopore. Data will be made available as a Zenodo repository.

## Funding

This study was funded by the UK Medical Research Council and the National Institute of Health Research (NIHR) Biomedical Research Centre at UCL Hospitals NHS Foundation Trust. S.M. and J.C. are NIHR Senior Investigators. F. K. is supported and funded by the Leonard Wolfson PhD Programme in Neurodegeneration. A.D. is supported by the Onassis Foundation - Scholarship ID: F ZQ 022-1/2020-2021.

## Authors’ contributions

S.M. had full access to all data in the study and takes responsibility for integrity of the data and accuracy of the data analysis. Concept and design: E.A.V. and S.M.; acquisition, analysis or interpretation of data: F.K., A.D., T.C., L.D., S.M. and E.A.V.; drafting of manuscript: F.K., A.D., E.A.V.; obtaining funding: J.C. and S.M.; study supervision: S.M. and E.A.V. All authors read and approved the final manuscript.

## Acknowledgments

We are grateful to patients and volunteers who have made this study possible by donating samples to research; to Paulo Amaral, Alejandro Gener (Baylor College of Medicine), Prof. Christos Proukakis (UCL) and Mike Clark (University of Melbourne) for helpful discussions; and to Joanna Field and Sarah Mazdon for help with metadata acquisition. The research was funded by the Medical Research Council (UK). Patient samples were provided with the assistance of funding from the UK’s National Institute of Health Research (NIHR) Biomedical Research Centre at University College London Hospitals NHS Foundation Trust. SM and JC are NIHR Senior Investigators.

## Competing interests

Prof. Collinge is a director and shareholder of D-Gen Limited (London), an academic spinout company working in the field of prion disease diagnosis, decontamination and therapeutics. François Kroll owns stocks in Oxford Nanopore Technologies and Pacific Biosciences. All other authors declare no competing interests.

## Notes

### Author Declarations

All experimental protocols were approved by the London Queen Square Research Ethics Committee (reference 05/Q0505/113).

## References

1. Mead S, Lloyd S, Collinge J. Genetic Factors in Mammalian Prion Diseases. Annu Rev Genet. 2019;53:117–47.

2. Oesch B, Westaway D, Walchli M, McKinley MP, Kent SB, Aebersold R, et al. A cellular gene encodes scrapie PrP 27-30 protein. Cell. 1985;40(4):735–46.

3. Yates AD, Achuthan P, Akanni W, Allen J, Allen J, Alvarez-Jarreta J, et al. Ensembl 2020. Nucleic Acids Res. 2020;48(D1):D682–D8.

4. Palmer MS, Dryden AJ, Hughes JT, Collinge J. Homozygous prion protein genotype predisposes to sporadic Creutzfeldt-Jakob disease. Nature. 1991;352(6333):340–2.

5. Pocchiari M, Puopolo M, Croes EA, Budka H, Gelpi E, Collins S, et al. Predictors of survival in sporadic Creutzfeldt-Jakob disease and other human transmissible spongiform encephalopathies. Brain. 2004;127(Pt 10):2348–59.

6. Goldfarb LG, Brown P, McCombie WR, Goldgaber D, Swergold GD, Wills PR, et al. Transmissible familial Creutzfeldt-Jakob disease associated with five, seven, and eight extra octapeptide coding repeats in the PRNP gene. Proc Natl Acad Sci U S A. 1991;88(23):10926–30.

7. Wadsworth JD, Joiner S, Linehan JM, Cooper S, Powell C, Mallinson G, et al. Phenotypic heterogeneity in inherited prion disease (P102L) is associated with differential propagation of protease-resistant wild-type and mutant prion protein. Brain. 2006;129(Pt 6):1557–69.

8. Li B, Qing L, Yan J, Kong Q. Instability of the octarepeat region of the human prion protein gene. PLoS One. 2011;6(10):e26635.

9. Mead S, Webb TE, Campbell TA, Beck J, Linehan JM, Rutherfoord S, et al. Inherited prion disease with 5-OPRI: phenotype modification by repeat length and codon 129. Neurology. 2007;69(8):730–8.

10. Bueler H, Aguzzi A, Sailer A, Greiner RA, Autenried P, Aguet M, et al. Mice devoid of PrP are resistant to scrapie. Cell. 1993;73(7):1339–47.

11. Westaway D, Mirenda CA, Foster D, Zebarjadian Y, Scott M, Torchia M, et al. Paradoxical shortening of scrapie incubation times by expression of prion protein transgenes derived from long incubation period mice. Neuron. 1991;7(1):59–68.

12. Vollmert C, Windl O, Xiang W, Rosenberger A, Zerr I, Wichmann HE, et al. Significant association of a M129V independent polymorphism in the 5’ UTR of the PRNP gene with sporadic Creutzfeldt-Jakob disease in a large German case-control study. J Med Genet. 2006;43(10):e53.

13. Jones E, Hummerich H, Vire E, Uphill J, Dimitriadis A, Speedy H, et al. Identification of novel risk loci and causal insights for sporadic Creutzfeldt-Jakob disease: a genome-wide association study. Lancet Neurol. 2020;19(10):840–8.

14. Sanchez-Juan P, Bishop MT, Croes EA, Knight RS, Will RG, van Duijn CM, et al. A polymorphism in the regulatory region of PRNP is associated with increased risk of sporadic Creutzfeldt-Jakob disease. BMC Med Genet. 2011;12:73.

15. Juling K, Schwarzenbacher H, Williams JL, Fries R. A major genetic component of BSE susceptibility. BMC Biol. 2006;4:33.

16. Hills D, Comincini S, Schlaepfer J, Dolf G, Ferretti L, Williams JL. Complete genomic sequence of the bovine prion gene (PRNP) and polymorphism in its promoter region. Anim Genet. 2001;32(4):231–2.

17. Sander P, Hamann H, Drogemuller C, Kashkevich K, Schiebel K, Leeb T. Bovine prion protein gene (PRNP) promoter polymorphisms modulate PRNP expression and may be responsible for differences in bovine spongiform encephalopathy susceptibility. J Biol Chem. 2005;280(45):37408–14.

18. De Coster W, De Rijk P, De Roeck A, De Pooter T, D’Hert S, Strazisar M, et al. Structural variants identified by Oxford Nanopore PromethION sequencing of the human genome. Genome Res. 2019;29(7):1178–87.

19. Beyter D, Ingimundardottir H, Oddsson A, Eggertsson HP, Bjornsson E, Jonsson H, et al. Long-read sequencing of 3,622 Icelanders provides insight into the role of structural variants in human diseases and other traits. Nat Genet. 2021;53(6):779–86.

20. Beck JA, Poulter M, Campbell TA, Adamson G, Uphill JB, Guerreiro R, et al. PRNP allelic series from 19 years of prion protein gene sequencing at the MRC Prion Unit. Hum Mutat. 2010;31(7):E1551–63.

21. Mahal SP, Asante EA, Antoniou M, Collinge J. Isolation and functional characterisation of the promoter region of the human prion protein gene. Gene. 2001;268(1-2):105-14.

22. Payne A, Holmes N, Rakyan V, Loose M. BulkVis: a graphical viewer for Oxford nanopore bulk FAST5 files. Bioinformatics. 2019;35(13):2193–8.

23. Beck JA, Poulter M, Campbell TA, Uphill JB, Adamson G, Geddes JF, et al. Somatic and germline mosaicism in sporadic early-onset Alzheimer’s disease. Hum Mol Genet. 2004;13(12):1219–24.

24. Kim YC, Won SY, Jeong BH. Identification of Prion Disease-Related Somatic Mutations in the Prion Protein Gene (PRNP) in Cancer Patients. Cells. 2020;9(6).

25. Leija-Salazar M, Piette C, Proukakis C. Review: Somatic mutations in neurodegeneration. Neuropathol Appl Neurobiol. 2018;44(3):267–85.

26. Park JS, Lee J, Jung ES, Kim MH, Kim IB, Son H, et al. Brain somatic mutations observed in Alzheimer’s disease associated with aging and dysregulation of tau phosphorylation. Nat Commun. 2019;10(1):3090.

27. White R PC, Ronchese F et al. Investigation of chimeric reads using the MinION. F1000Research. 2017;6:631.

28. Xu Y, Lewandowski K, Lumley S, Pullan S, Vipond R, Carroll M, et al. Detection of Viral Pathogens With Multiplex Nanopore MinION Sequencing: Be Careful With Cross-Talk. Front Microbiol. 2018;9:2225.

29. Delahaye C, Nicolas J. Sequencing DNA with nanopores: Troubles and biases. PLoS One. 2021;16(10):e0257521.

30. Gilpatrick T, Lee I, Graham JE, Raimondeau E, Bowen R, Heron A, et al. Targeted nanopore sequencing with Cas9-guided adapter ligation. Nat Biotechnol. 2020;38(4):433–8.

31. I S. Comprehensive genetic diagnosis of tandem repeat expansion disorders with programmable targeted nanopore sequencing. medRxiv. 2021.

32. Guere ME, Vage J, Tharaldsen H, Benestad SL, Vikoren T, Madslien K, et al. Chronic wasting disease associated with prion protein gene (PRNP) variation in Norwegian wild reindeer (Rangifer tarandus). Prion. 2020;14(1):1–10.

33. Nonno R, Di Bari MA, Pirisinu L, D’Agostino C, Vanni I, Chiappini B, et al. Studies in bank voles reveal strain differences between chronic wasting disease prions from Norway and North America. Proc Natl Acad Sci U S A. 2020;117(49):31417–26.

34. Lopez Castel A, Cleary JD, Pearson CE. Repeat instability as the basis for human diseases and as a potential target for therapy. Nat Rev Mol Cell Biol. 2010;11(3):165–70.

35. Origin of extra prion repeat units. 1999.

36. Metzgar D, Liu L, Hansen C, Dybvig K, Wills C. Domain-level differences in microsatellite distribution and content result from different relative rates of insertion and deletion mutations. Genome Res. 2002;12(3):408–13.

37. Gomes-Pereira M, Fortune MT, Ingram L, McAbney JP, Monckton DG. Pms2 is a genetic enhancer of trinucleotide CAG.CTG repeat somatic mosaicism: implications for the mechanism of triplet repeat expansion. Hum Mol Genet. 2004;13(16):1815–25.

38. Monckton DG. The Contribution of Somatic Expansion of the CAG Repeat to Symptomatic Development in Huntington’s Disease: A Historical Perspective. J Huntingtons Dis. 2021;10(1):7–33.

39. Eid J, Fehr A, Gray J, Luong K, Lyle J, Otto G, et al. Real-time DNA sequencing from single polymerase molecules. Science. 2009;323(5910):133–8.

40. Dabin LC, Guntoro F, Campbell T, Belicard T, Smith AR, Smith RG, et al. Altered DNA methylation profiles in blood from patients with sporadic Creutzfeldt-Jakob disease. Acta Neuropathol. 2020;140(6):863–79.

41. Beck JA, Mead S, Campbell TA, Dickinson A, Wientjens DP, Croes EA, et al. Two-octapeptide repeat deletion of prion protein associated with rapidly progressive dementia. Neurology. 2001;57(2):354–6.

42. Koren S, Walenz BP, Berlin K, Miller JR, Bergman NH, Phillippy AM. Canu: scalable and accurate long-read assembly via adaptive k-mer weighting and repeat separation. Genome Res. 2017;27(5):722–36.

43. J S. Nanopolish. 2020.

44. Li H. Minimap2: pairwise alignment for nucleotide sequences. Bioinformatics. 2018;34(18):3094–100.

45. Li H, Handsaker B, Wysoker A, Fennell T, Ruan J, Homer N, et al. The Sequence Alignment/Map format and SAMtools. Bioinformatics. 2009;25(16):2078–9.

46. GATKTeam. StrandOddsRatio. 2019.

47. M M. WhatsHap: fast and accurate read-based phasing. bioRxiv. 2016.

48. Sedlazeck FJ, Rescheneder P, Smolka M, Fang H, Nattestad M, von Haeseler A, et al. Accurate detection of complex structural variations using single-molecule sequencing. Nat Methods. 2018;15(6):461–8.

49. Thompson JD, Higgins DG, Gibson TJ. CLUSTAL W: improving the sensitivity of progressive multiple sequence alignment through sequence weighting, position-specific gap penalties and weight matrix choice. Nucleic Acids Res. 1994;22(22):4673–80.

50. R package for multiple sequence alignment.

51. Piovesan A, Pelleri MC, Antonaros F, Strippoli P, Caracausi M, Vitale L. On the length, weight and GC content of the human genome. BMC Res Notes. 2019;12(1):106.

