## Supplementary for "Prion protein gene mutation detection using long-read Nanopore sequencing"

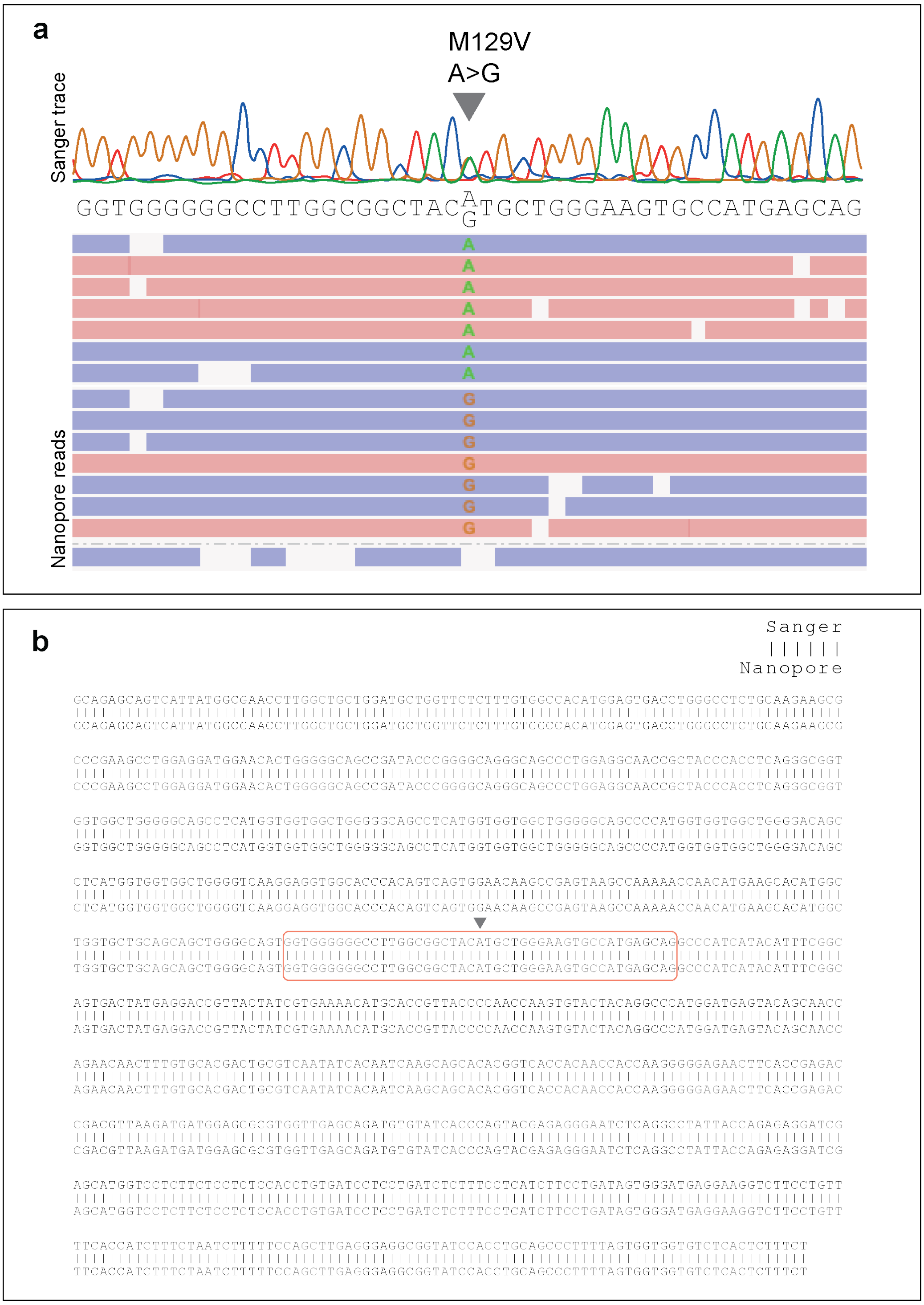


**Figure S1. Sequencing of *PRNP*'s protein-coding region using Oxford Nanopore MinION**

(**a**) Genomic DNA from one patient's blood sample was genotyped as codon 129 heterozygous (arrowhead) using Sanger sequencing. The SNV was specifically detected from the Nanopore reads at the correct zygosity. (**b**) Alignment of the sequence obtained from Sanger sequencing to the consensus sequence computed from the Nanopore reads. The orange frame indicates the region depicted in (a), the arrowhead indicates the codon 129 SNV.

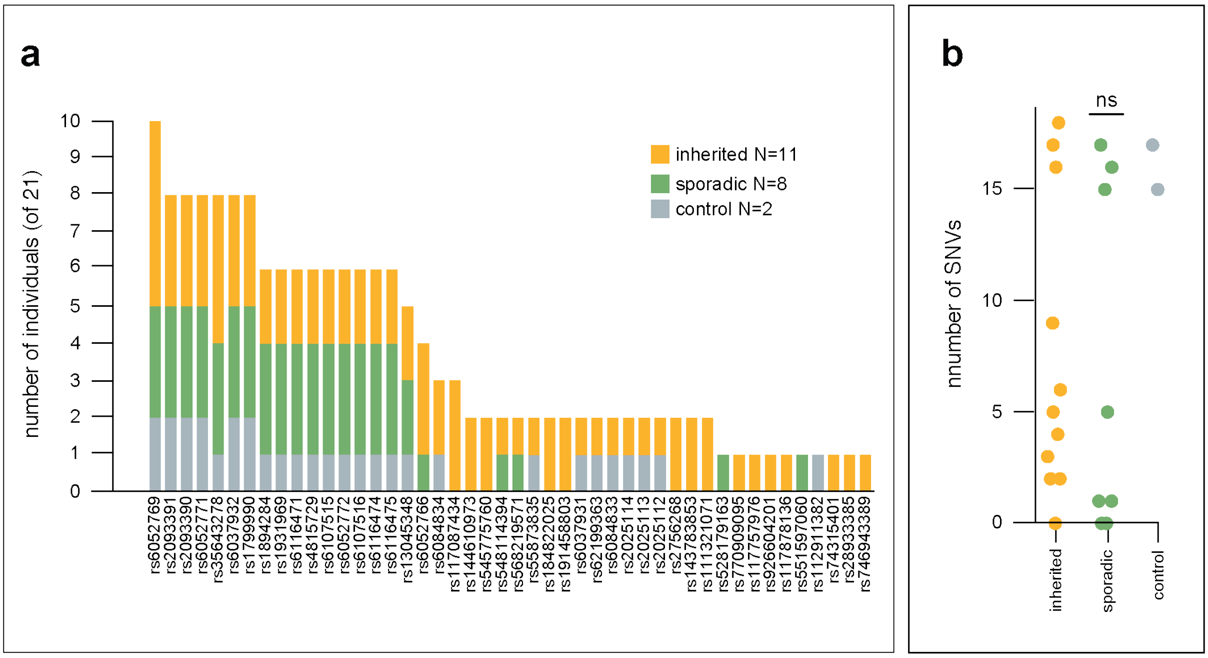


**Figure S2. Detection of SNVs in PRNP with Nanopore sequencing**

(**a**) Number of individuals from our panel who carried each SNV. (**b**) Number of SNVs carried by each individual, grouped by cohort. Each dot represents one individual. Individuals from different cohorts did not carry significantly different numbers of SNVs. ns, $p = 0.24$ by one-way ANOVA.

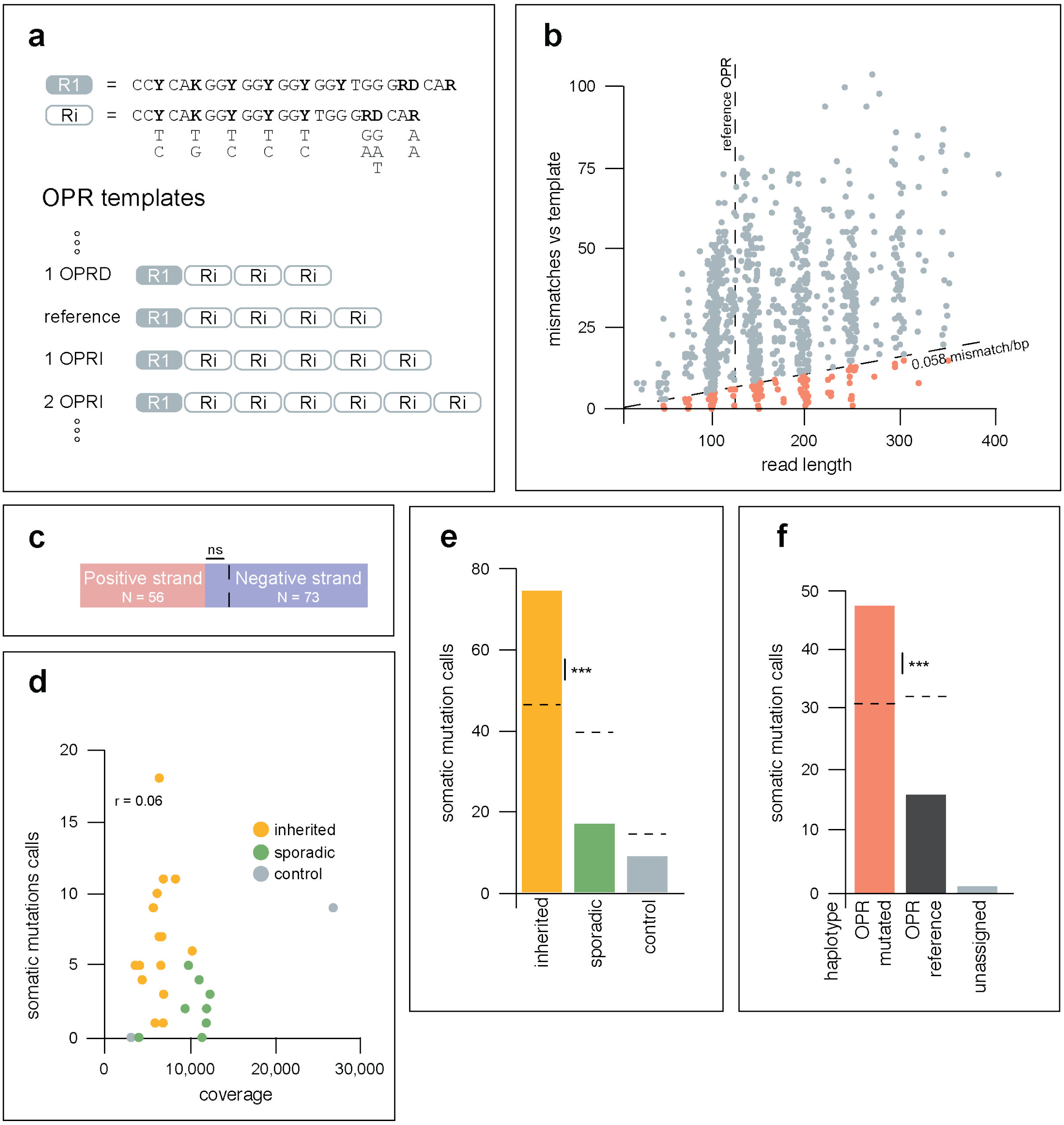


**Figure S3. Detection of candidate somatic mutations in *PRNP*'s OPR.**

(**a**) Ri is a consensus sequence for one octapeptide repeat. It uses flexible nucleotide codes (e.g. Y is either T or C) to allow R2, R3, R4 and other possible variants reported in CJD cases. We then used Ri to build templates for a range of possible OPRs, from deletion of 4 repeats (only R1 remaining) up to insertion of 24 repeats by adding Ri sequences. (**b**) Each dot represents one candidate somatic mutation read aligned to its most likely OPR template. The vertical dashed line marks the length of the reference OPR at 123 bp. The black diagonal line marks the mismatch threshold (0.058 mismatch/bp, i.e. at least 94.2% of the read must align to its OPR template). Reads below the threshold were selected as somatic mutations calls (N = 129, orange). (**c**) No evidence of strand bias in the somatic mutation calls. Of the N = 129 somatic mutation calls, N = 56 were from the positive strand (pink) and N = 73 were from the negative strand (blue). This was not significantly different than the expected counts (62.2 and 66.8 from positive and negative strand, respectively; dashed black line). ns, p = 0.27 by Pearson's Chi-squared test. (**d**) Number of somatic mutation calls per sample did not correlate with sequencing coverage. r = 0.06 by Pearson's correlation. (**e**) Total number of somatic mutation calls from each group of samples (inherited CJD, sporadic CJD, controls). Of the N = 101 somatic mutation calls from blood samples, N = 75 were from inherited CJD samples, N = 17 were from sporadic CJD samples, and N = 9 were from control samples. There was a surplus of somatic mutation calls from inherited CJD samples. *** p < 0.001 by Pearson's Chi-squared test (expected counts: 37.6, 46.5, 16.9 from inherited CJD, sporadic CJD, and control samples respectively, dashed black lines). (**f**) Total number of somatic mutation calls from each haplotype. Only samples which could be haplotype-phased and had an OPR mutation were included here. Of N = 64 somatic mutation calls from these samples, N = 47 were in reads from the haplotype carrying the OPR mutation, N = 16 were in reads from the haplotype carrying the reference OPR, N = 1 was on a read which could not be assigned to a haplotype. There was a surplus of somatic mutation calls in reads from the mutated OPR haplotype. *** p < 0.001 by Pearson's Chi-squared test (expected counts: 30.9 and 32.0 from the mutated haplotype and the reference haplotype respectively; dashed black lines).

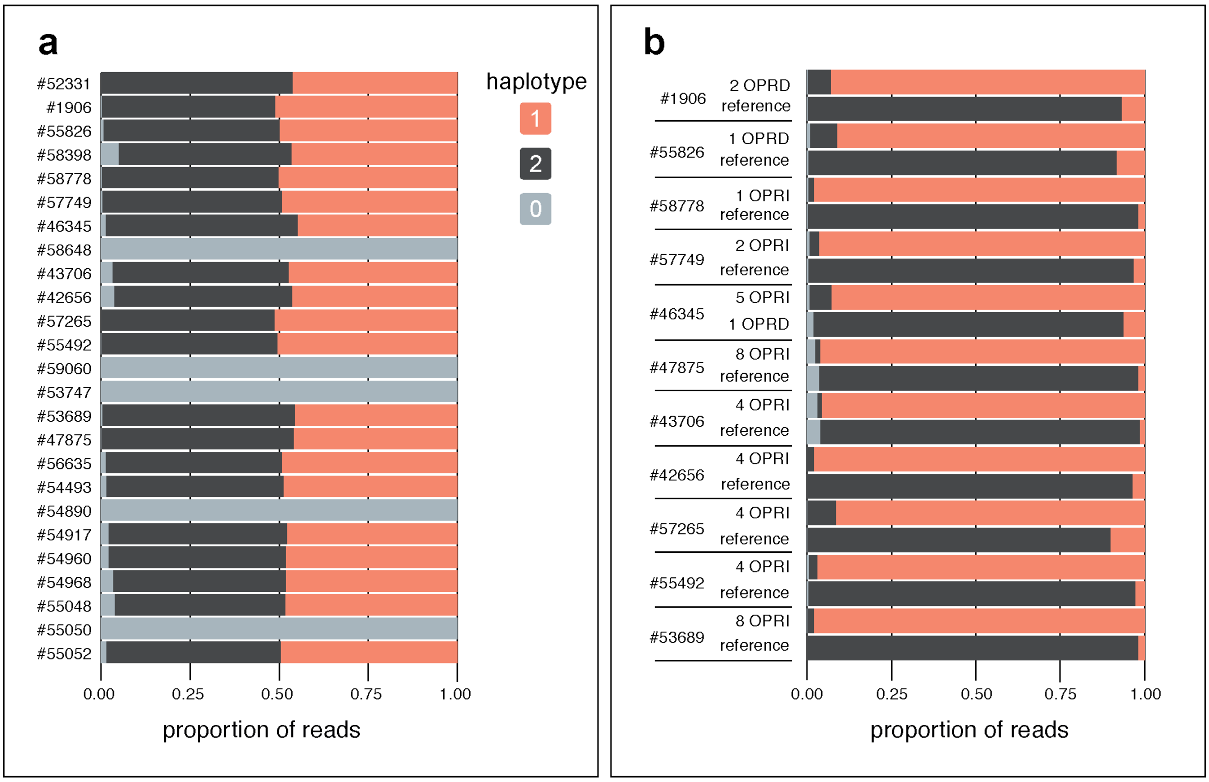


**Figure S4. Assigning reads to haplotypes**

(**a**) For each sample, proportion of reads assigned to haplotype 1 (orange) or haplotype 2 (black) or unassigned (haplotype 0, grey). The samples for which all the reads were unassigned did not have any heterozygous SNV which could be used for haplotype phasing (**b**) For each sample, haplotype assignments of the reads carrying the mutated OPR or carrying the reference OPR. Only samples which could be haplotype-phased and had a OPR mutation were included here. For these samples, haplotype 1 corresponds to the mutated OPR haplotype, haplotype 2 corresponds to the reference OPR haplotype.

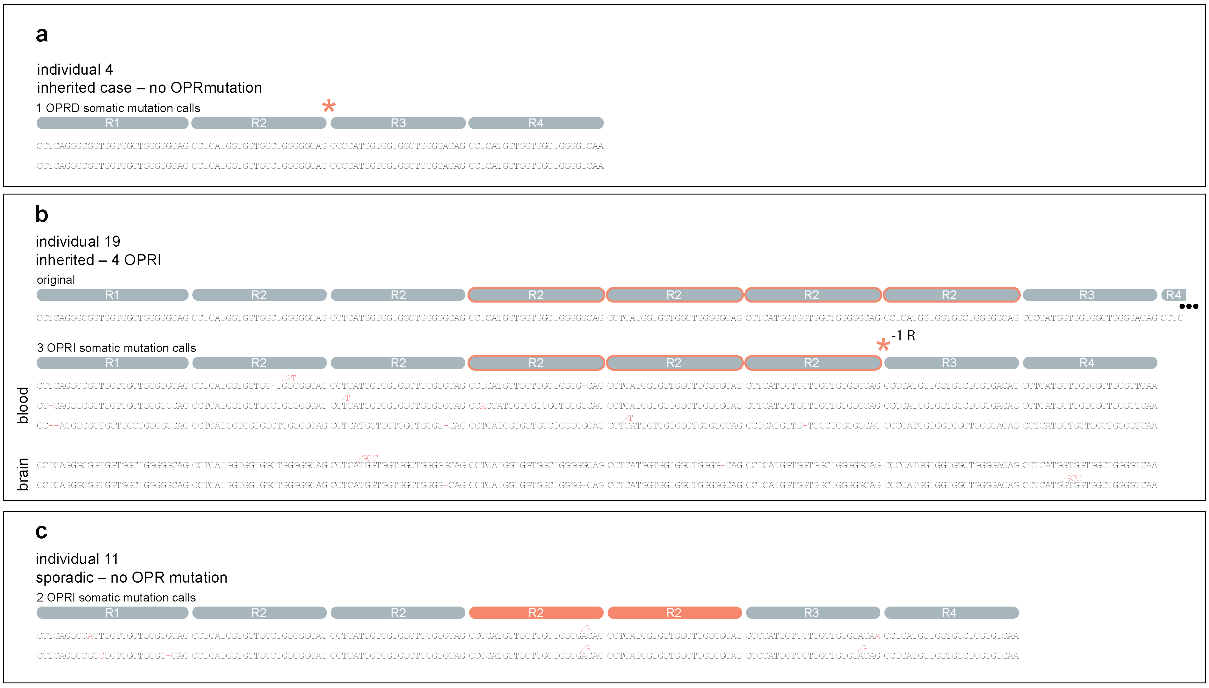


**Figure S5. Examples of somatic mutation calls**

(**a**) Individual 4 is an inherited CJD case with reference OPRs. The two reads shown are 1 OPRD calls (R2 deletion) with 0 mismatch with the template. (**b**) Individual 19 is an inherited CJD case with a heterozygous 4 OPRI mutation. The top read (original) represents the 4 OPRI mutation. The next three reads are examples of 3 OPRI somatic mutation calls (likely R2 deletion) from the blood sample of this individual, the next two reads are examples of 3 OPRI somatic mutation calls (likely R2 deletion) from the brain sample of the same individual. Small, likely artefactual, indels are represented by red dashes (deletions) or red nucleotides on top of the read (insertions). (**c**) Individual 11 is a sporadic CJD case with reference OPRs. The two reads shown are 2 OPRI somatic mutation calls.

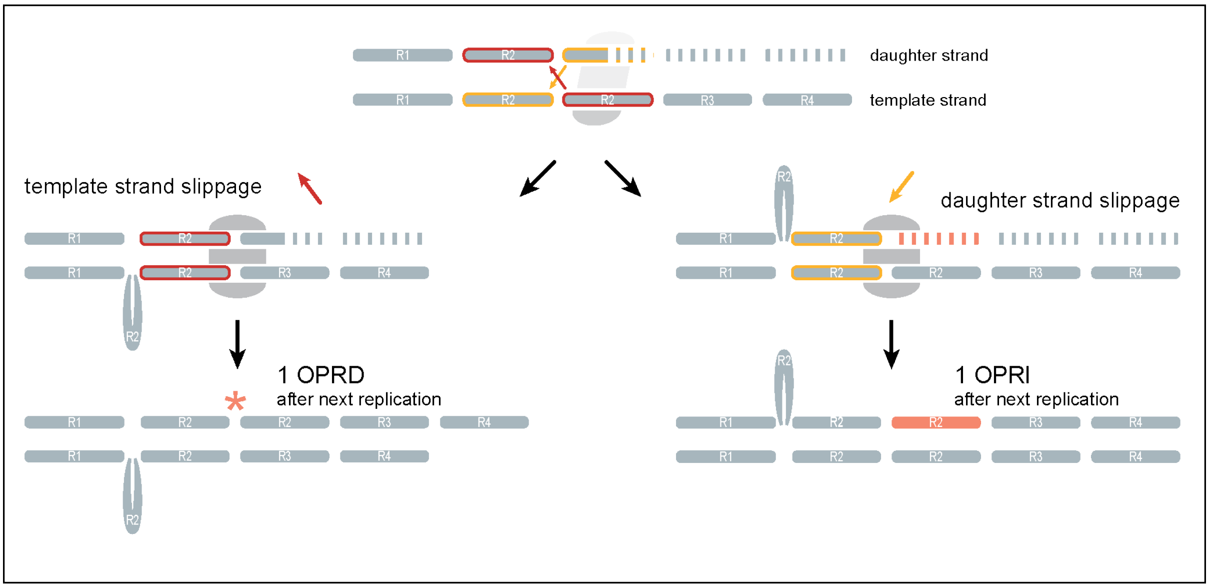


**Figure S6. Possible generation of OPR mutations by replication slippage**

(**a**) Mechanism by which replication slippage may generate OPR mutations. If the DNA polymerase falls off while replicating *PRNP*'s OPR, the template and daughter strands may re-anneal incorrectly due to the repeats. During template strand slippage (left), a repeat in the template strand incorrectly anneals with an earlier repeat in the daughter strand. For example, the second R2 repeat may wrongly anneal with the newly synthesised first R2 in the daughter strand (red arrow). This results in a repeat extruding from the template strand. When the DNA polymerase resumes replication, it misses one or more repeat(s), generating an OPR deletion in the daughter strand. The result is initially an heteroduplex molecule, which becomes an OPR deletion on both strands during the next replication when the strand with an OPR deletion becomes the template strand. Similarly, during daughter strand slippage (right), a repeat in the daughter strand incorrectly anneals with an earlier repeat in the template strand. For example, the newly synthetised second R2 may wrongly anneal with the first R2 in the template strand (yellow arrow). This results in a repeat extruding from the daughter strand. When the DNA polymerase resumes replication, it replicates the same repeat(s) again, generating an OPR insertion in the daughter strand. The result is initially an heteroduplex molecule, which becomes an OPR insertion on both strands during the next replication when the strand with an OPR insertion becomes the template strand. The mechanism is depicted here as respecting repeat boundaries only for simplicity. In practice, there are many ways the repeats can misalign by multiples of 24 bp.
